# Disentangling the link between maternal influences on birth weight and disease risk in 36,211 genotyped mother-child pairs

**DOI:** 10.1101/2023.10.17.23297154

**Authors:** Jaakko T. Leinonen, FinnGen, Matti Pirinen, Taru Tukiainen

## Abstract

Epidemiological studies have robustly linked lower birth weight to later-life disease risks. These observations may reflect the adverse impact of intrauterine growth restriction on a child’s health. However, causal evidence supporting such a mechanism in humans is largely lacking.

Using Mendelian Randomization and 36,211 genotyped mother-child pairs from the FinnGen study, we assessed the relationship between intrauterine growth restriction and five common health outcomes (coronary heart disease (CHD), hypertension, statin use, type 2 diabetes and cancer). We proxied intrauterine growth restriction with polygenic scores for maternal effects on birth weight and took into account transmission of genetic variants between a mother and a child in the analyses.

We find limited evidence for contribution of normal variation in maternally influenced intrauterine growth on later-life disease. Instead, we find support for genetic pleiotropy in the child genome linking birth weight to CHD and hypertension. Our study illustrates the opportunities that data from genotyped parent-child pairs from a population-based biobank provides for addressing causality of maternal influences.

## Introduction

The relationship between birth weight and disease risk has been extensively studied in epidemiological settings, revealing associations with adverse health consequences both for lower and higher birth weights ^1–8^. Of particular note is the connection between low birth weight and cardiometabolic diseases ^9,10^. This connection has been speculated to arise from long-lasting changes in metabolic programming due to intrauterine growth restriction (IUGR), referring to poor fetal growth ^9,10^. This concept has led to the hypothesis of developmental origins of health and disease (DOHaD) suggesting that several non-communicable diseases originate in early development - during prenatal life, in an unfavourable intrauterine environment, or in early childhood ^11^. The causal evidence in support of the DOHaD mechanism in humans has been, however, largely lacking and conflicting ^3,12,13^. Consequently, although the relationship between birth weight and disease risk is clear at the epidemiological level, the underlying causes behind these associations have been a subject for debate for decades ^3^.

Mendelian randomization (MR) has become a popular method to assess causal relationships between an exposure and an outcome ^14^. In short, MR utilizes genetic variants robustly associated with the exposure to test whether the same variants have consistent effects on the outcome. MR strategies using genetic variants of the mother associated with birth weight of the child therefore offer a means to assess the potential causal influences of intrauterine growth restriction as approximated by the birth weight of the child ^12,14,15^.

Variation in birth weight has a large genetic component (SNP-based heritability (h^2^_SNP_ being ∼40%, of which a fifth can be specifically allocated to maternal genetic variation), making it an amenable trait for genetic studies ^15^. However, since many different factors including both the maternal and fetal genome influence the size of the baby, using MR in assessing the DOHaD hypothesis has some important prerequisites ^3,15–18^. First, using genetic factors specifically reflecting maternal influences on birth weight as instrumental variables in MR is crucial ^12,19^, as these can plausibly reflect intrauterine growth restriction as opposed to the variants with strict fetal effects. Secondly, a standard two sample MR using the maternal genotype only can lead to biased causality estimates as it fails to account for the 50% correlation between the maternal and fetal genotypes, therefore violating the exclusivity assumption of MR ^12,20^. To overcome this issue, data sets with genotyped mother-child pairs are of great value since these allow for blocking the path through the child’s genome by adjusting the analysis with information on child’s genetic variants at the loci tested.

Recent studies utilizing these MR principles and mother-child pairs suggest that small genetic effects on IUGR have only limited effects on child’s cardiometabolic risk factors ^12,15^. These studies, based on mother-child pairs from a Norwegian HUNT study (N=26,057) ^12^ and UK Biobank (N=3,886) ^15^, have reported lack of strong effects of IUGR on cardiometabolic risk factors such as lipid and glucose levels and hypertension. To our knowledge, comparable studies are limited, primarily due to the scarcity of suitable genotyped parent-child cohorts ^21^. Additionally, it remains uncertain whether these results might extend to the development of cardiovascular disease.

Population-based biobank data may offer new possibilities to conduct MR studies of maternal exposures ^12,22^, as these data typically capture also familial relationships due their large scale of sampling. For instance, within the 430,897 participants of the FinnGen study (release 10) ^23^, corresponding to 8% of the Finnish population, there are 67,986 first-degree relationships including 36,211 mother-child and 31,775 father-child pairs with genotype and extensive longitudinal health registry information available. A particular advantage of the parent-child pairs contained within FinnGen is the comparatively advanced age of the children (mean 46.3 yrs., SD 14.2), allowing for examination of disease endpoints that manifest later in life.

Here, we apply an established MR framework ^12^ to 36,211 mother-child pairs from the FinnGen study to examine how polygenic scores (PGS) for maternal influences on birth weight associate with five common health outcomes in the offspring (Figure 1; N cases = 996 - 6,150), accounting for the effects from both the maternal and offspring genome in the same model. Extending the previous findings on biomarkers^12,15^, we find no evidence for the role of IUGR, as proxied by genetic scores for maternal effects on birth weight, in determining child’s disease risks. Rather, we show that the scores of the same variants in the fetal genome associate with CHD and hypertension, an effect that is also detected in the 31,775 father-child pairs from FinnGen. Together these findings suggest that genetic pleiotropy in the child is largely accountable for the epidemiological links between birth weight and disease.

**Figure 1.**
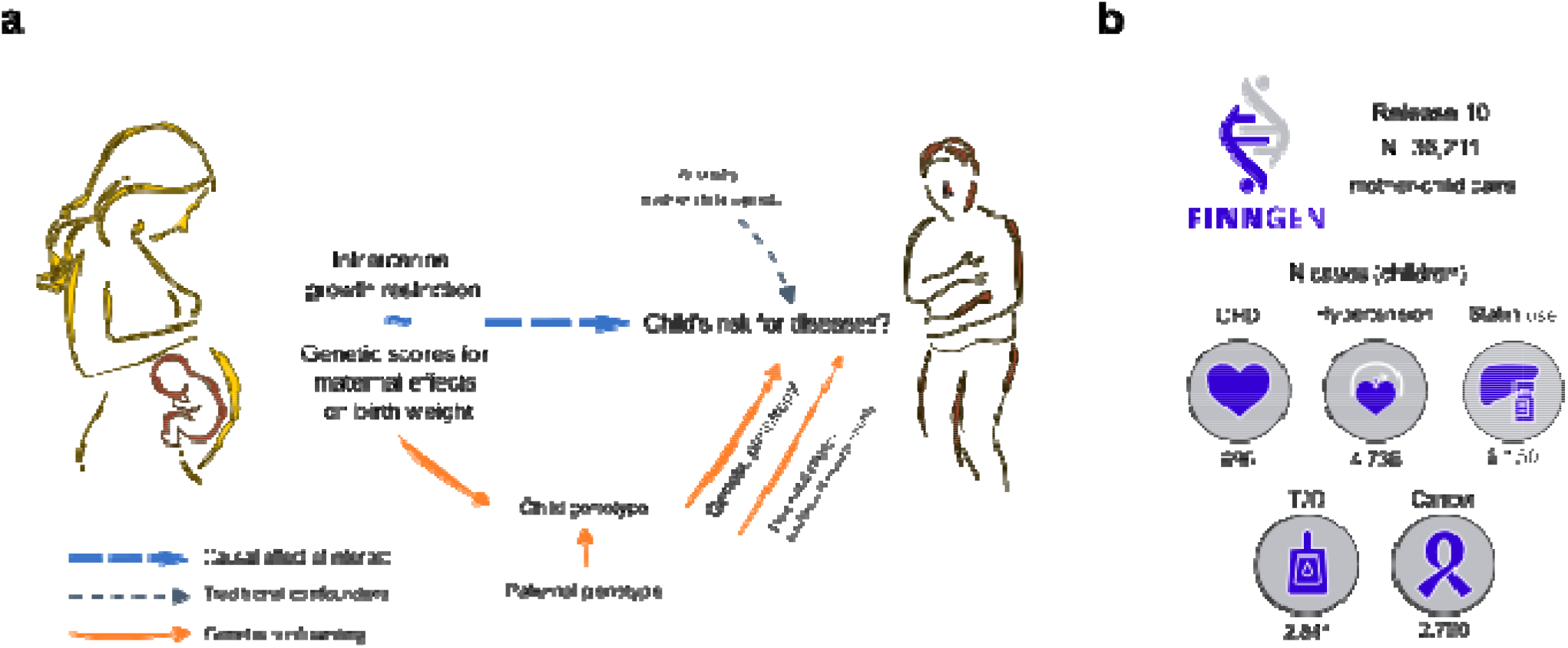
Key aspects of the study. A) Illustration of the Mendelian Randomisation (MR) framework to assess relationship between intrauterine growth restriction (IUGR) and child’s disease risk when IUGR is approximated by child’s birthweight. This MR analysis tests whether mother’s genetic score for child’s birth weight (M-GS) is associated with child’s disease risk later in life. MR relies on a principle that M-GS does not affect child’s disease risk through any other pathway than IUGR. Since the child inherits 50% of genetic variants from the mother, also roughly half of the variants contributing to M-GS are transmitted to the child. Theoretically, the same variants which in the mother affect child’s size can have other functions when transmitted to the child (genetic pleiotropy). Therefore, it is vital to account for this genetic sharing by conditioning the MR analyses on the child’s genetic score (C-GS) for the same variants that are used in M-GS. b) Summary information of the FinnGen data set with the case numbers in the children from the mother-child pairs indicated for each disease. CHD = coronary heart disease, T2D = Type 2 Diabetes.

## Results

### Polygenic scores predict birth weight in FinnGen

We first constructed polygenic scores (PGSs) for birth weight using the findings of the latest genome-wide association study (GWAS) of person’s own (n=321,223) and offspring birth weight (n=230,069 mothers) that applied structural equation modelling to dissect the birth weight associated genetic markers (N total = 209) into those with maternal only, fetal only, or shared effects^15^. Using these GWAS results, we built four PGSs that capture a different degree of maternal influences on child’s birth weight. To exclusively model the maternal contribution to birth weight, we used two PGSs reflecting strictly maternal effects on birth weight: a score based on 29 lead SNPs with only maternal effects on birth weight (MBW-29) and a respective genome-wide score (MBW-GW). We supplemented these with two other lead SNP-based scores (BW-68 and BW-201) that contain variants with both maternal and fetal effects on birth weight (Methods and Supplementary Figure 1).

Since we aimed to employ these PGSs as instrumental variables within a Mendelian Randomization (MR) framework to assess causality, our first step was to confirm that the PGSs predict measured birth weight in FinnGen. To this end we utilized a subset of FinnGen participants with birth weight measurements available, based on national birth registry (FinnGen R10 N = 39,578; mother-child pairs N = 9,257; father-child pairs N = 5,740).

All birth weight PGSs had statistically significant effects on birth weight when computed from an individual’s own genotypes in the full FinnGen data (p < 0.05; Supplementary Table 1 and Supplementary Figure 2). In mother-child pairs, BW-68 and BW-201 PGSs showed associations to both child and own birth weight, as expected since these scores contain variants with both maternal and fetal effects on birth weight. Importantly, however, the two PGSs based on variants with maternal effects, MBW-29 and MBW-GW, showed mother-specific effects on child’s birth weight. In other words, here only the mother’s score influenced child’s birth weight (at significance level p < 0.05) when including both mother’s and child’s PGSs to a multiple regression model (Supplementary Table 1).

To further validate the predictive values of the PGSs, we computed them for the fathers and children of father-child pairs from FinnGen. Here, as expected given these scores should tag maternal effects, neither MBW-29 nor MBW-GW of the father associated with child’s birth weight (Supplementary Table 1). However, for MBW-GW we noted a significant negative association (p < 0.05) of father’s PGSs on child’s birth weight after including the child’s PGS in the model ^24,25^. This can indicate both the presence of a potential collider bias and the fact that a genome-wide score may contain an excess of variants that are not specific to the trait studied (genetic pleiotropy). We therefore chose to focus on MBW-29 as our main instrument for further analyses.

### Association of maternal birth weight polygenic scores with child’s disease risk

The connections between birth weight and several diseases are well established at both epidemiological and genetic level ^1,4–7,12,15^, and echoing these observations we detected many associations between birth weight PGSs computed from person’s own genotypes and disease outcomes in the FinnGen dataset (N=412,176) (Supplementary Table 2). While these population-level analyses indicate clear relationships between lower birth weight and higher risk, e.g., for cardiovascular disease, these do not allow the assessment of DOHaD as the underlying causal mechanism as the influence of the birth weight variants are examined in one’s own genome.

To address potential causality of DOHaD, we therefore focused on understanding how the PGSs specific for maternal effects on birth weight, proxying intrauterine growth restriction, associate with disease risk in the 36,211 mother-child pairs in FinnGen. A key component of the MR framework applied here is to include both the mother’s and the child’s PGS to the analyses to control for the 50% correlation between these PGSs (Figure 1 and Supplementary Figure 1) ^12^. A failure to adjust the analyses with the child’s own PGS can lead to a spurious association between maternal PGS and child’s diseases, as shown by our simulations (Supplementary Table 3 and Supplementary Figure 3). Instead, in case of true intrauterine effects, we would expect that any association between a mother’s PGS and child’s disease risk remains unchanged when adding information from the child’s PGS to the model (Supplementary Table 3).

When analysing the effects of the mother’s and child’s PGS separately (i.e., including only one of the PGSs into the model) both the maternal and child’s own PGS showed several statistically significant associations with different diseases (p < 0.05). For example, higher mother’s MBW-29 PGS was associated with a reduced risk for CHD (OR=0.92, [95% CI 0.86-0.97], p=0.0089) and statin use, (OR=0.96 [95% CI 0.93-0.99], p=0.016), and increased risk for cancer (1.04 [95% CI 1.00-1.08], p=0.049). Child’s own PGS was similarly associated with these disease outcomes (for MBW-29 OR=0.87 [95% CI 0.81-0.92], p=1.49e-05 (CHD); OR=0.96, [95% CI 0.93-0.99] p=0.024 (Hypertension); OR=0.97, [95% CI 0.94-0.99], p=0.022 (Statin use) and OR=1.05, [95% CI 1.01-1.09], p=0.015 (Cancer) (Figure 2, Supplementary Figure 3, and Supplementary Table 4).

**Figure 2.**
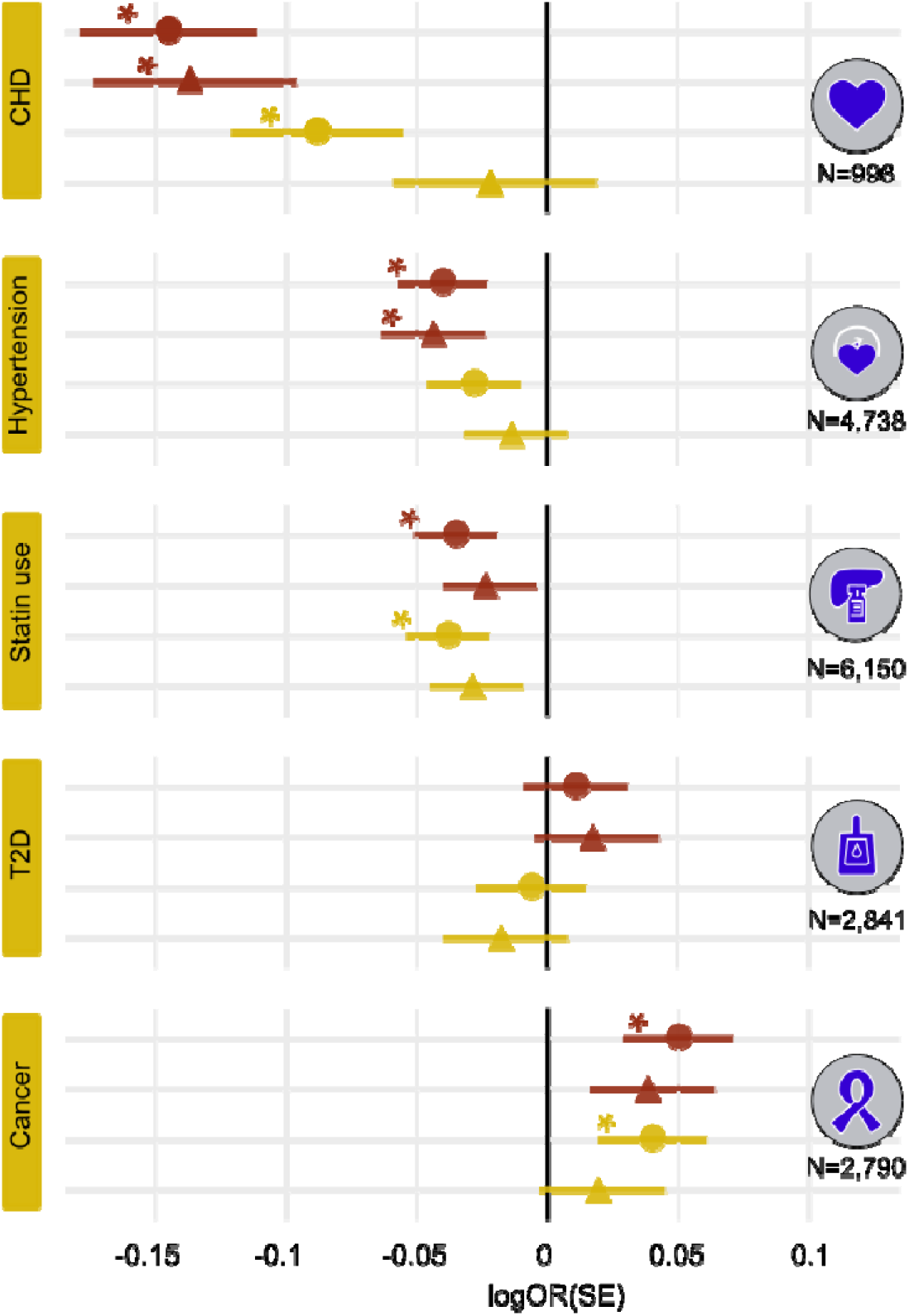
Associations of a PGS for maternal effects on birth weight (MBW-29) with child’s disease risk from mother-child pairs (N=36,211). The figure shows how the child’s own PGS (red) and mother’s PGS (yellow) associate with child’s disease risk in the mother-child pairs. Round dots illustrate the effect estimates (in logOR units) from analyses where only one PGS (either maternal or fetal MBW-29) is used, and triangels indicate that the analyses include MBW-29 PGSs from both mothers and their children. The lines around the effect estimates mark standard error (SE). * denotes statistically significant association (p < 0.05) between a PGS and disease risk in children.

However, when taking into account both mother’s and child’s PGS in the analyses simultaneously, the maternal PGS, that explained child’s birth weight irrespective of child’s PGS, no longer displayed associations with child’s disease risks (p > 0.05) (Figure 2, Supplementary Figure 3, and Supplementary Table 4). Here, matching the expectations from simulations, in two instances the effect sizes of the maternal MBW-29 PGS were reduced compared to the unadjusted models (from 0.97 to 0.99 for hypertension, and 0.92 to 0.98 for CHD) (Figure 2, Supplementary Figure 3, and Supplementary Table 4). Instead, in the same analyses child’s own MBW-29 PGS remained statistically significantly associated with both diseases, with comparable effect sizes as in the unadjusted analyses OR=0.87 [95% CI 0.81-0.94], p=0.00047 for CHD and OR=0.96 [95% CI 0.92-0.99], p=0.032 for hypertension).

While the results for CHD and hypertension aligned with the expectations of exclusive fetal effects (Supplementary Figures 2 and 3), for the three other traits the patterns of associations were complex, though non-significant in models including both maternal and child PGS (Figure 2, Supplementary Figure 3, and Supplementary Table 4). In case of statin use and cancer we observed significant associations (p < 0.05) in the models including only mother’s or child’s PGS. Yet, these became non-significant in the combined model, which may reflect reduced statistal power or may indicate joint maternal and fetal effects on these outcomes. For T2D, the maternal and fetal point estimates from the joint model were to the opposite directions, though with large standard errors (Figure 2, Supplementary Figure 3 and Supplementary Table 4).

### Sensitivity analyses

After these main analyses, we ran additional tests to probe the robustness of our findings. Although our focus was on the action of an unweighed lead-SNP based PGS specifically tagging maternal effects on birth weight (MBW-29), results from the other PGSs (MBW-GW, BW-68 and BW 205) supported the concept that birth weight – disease associations are largely driven by effects of the genetic variants in the child’s genome (Supplementary Figure 3 and Supplementary Table 4). None of the mother’s PGSs were associated with disease risk in the children after taking into account child’s own PGS (p > 0.05), and the maternal effect sizes were usually reduced as expected based on the simulations where the child’s own genome conferred the risk (Supplementary Figures 2 and 3, and Supplementary Tables 3 and 4).

We then noted that the lack of significant maternal effects held true notwithstanding whether we limited our dataset to include only one child per mother (N=28,582), or used a stringently filtered dataset where only a maximum of 4^th^ degree relatives from both mothers and children were included (N=22,454, Supplementary Table 5). Similarly, adjusting the results with a PGS for gestational duration did not support the presence of maternal effects (Supplementary Table 5). The only exception was statin use for which we detected nominal support for maternal effects in the stringently unrelated mother-child pairs (Supplementary Table 5). Importantly, we detected that the effect size for a mother’s PGSs did not change in a statistically significant manner in any of these sensitivity analyses (Supplementary Figure 4).

We finally followed up these findings in the 31,775 father-child pairs (child mean age 47.6 yrs. [SD 13.8]) available in FinnGen. Here, similarly as in the birth weight prediction (Supplementary Table 1), association of a father’s PGS with child’s disease risks would be unexpected, as no intrauterine mechanisms are in play, and any association would thus point to effects from the postnatal environment or confounding by assortative mating. Yet we would expect any effects of the fetal genome to be present also in the father-child pairs. In line with these expectations, we detected little evidence for any paternal effects, with the exception of paternal MBW-GW associating with statin use (Supplementary Figure 5 and Supplementary Table 6). Rather we repeatedly associated child’s own genetic scores for maternal effects on birth weight (MBW-29) with disease risk also in the father-child pairs, with comparable effect estimates as in mother-child pairs, e.g., OR=0.95 [95% CI 0.91-0.99], p=0.012 for hypertension and OR = 0.91 [95% CI 0.84-0.99], p=0.021 for CHD (Supplementary Figure 5 and Supplementary Table 6).

### Power Calculations

We conducted simulations to assess the magnitude of maternal and fetal effects we were sufficiently powered to detect in the 36,211 mother-child pairs for each endpoint (see Methods; Supplementary Table 3). These analyses indicated that when including both maternal and child PGSs in the analysis we had adequate power (≥80%) to detect true maternal effects (OR per 1SD change in PGS) ranging from >±1.11 for CHD to >±1.05 for statin use (Supplementary Table 3 and Supplementary Figure 6).

## Discussion

The relationship between birth weight and disease risks has been intensively studied for several decades, and many different theories have been proposed to explain their connections. In this study, we set out to examine the causal mechanisms between lower birth weight and later-life health using large-scale genetic data. In particular, we aimed to understand the proposed role of intrauterine growth restriction, i.e., the mechanism of the DOHaD hypothesis, as a determinant of the offspring disease risks. To this end, we used a specifically designed Mendelian randomization (MR) framework in a large sample of Finnish genotyped mother-child pairs. This framework overcomes many limitations of regular MR related to assessing maternal influences, allowing for more accurate estimation of causal effects of modest intrauterine growth restriction on child’s risk for diseases ^12^. Here intrauterine conditions are proxied using genetic markers with strict maternal effects on child’s birth weight, and the independent contribution of the maternal genome is assessed by blocking the direct transmission of alleles by adjusting the analysis using the child’s genetic score at the same loci.

Taking advantage of a unique data set of 36,211 genotyped mother-child pairs available within the FinnGen study, we applied the MR framework on five health registry endpoints epidemiologically associated with birth weight. Overall, we did not find support for a strong connection between intrauterine growth restriction and offspring later life disease. Rather, it seems that the same genetic variants that when present in mothers affect child’s birth weight, once inherited by the child have independed effects on child’s disease risks. Our findings thus point to the links of lower birth weight and diseases occuring largely due to genetic pleiotropy in the child’s genome.

Our study builds upon previous research investigating the DOHaD mechanisms using mother-child and biomarker data from biobanks, including the UK Biobank (N=3,886) ^15^ and the HUNT cohort (N=26,057) ^12^. In this study, we expand upon these analyses by using a larger number of mother-child pairs and examining the associations between maternal effects on birth weight and five binary disease outcomes sourced from nationwide health registries. Our decision to focus on binary outcomes, while potentially reducing statistical power, uniquely positions us to directly explore the link between genetically determined maternal effects on birth weight and disease manifestations, moving beyond the previous investigations of disease risk factors.

Given the lack of statistically significant maternal contribution to child’s disease risks, our results suggest that modest intrauterine growth restriction may have limited effects on diseases such as CHD and hypertension compared to the effects of child’s own genome. Echoing the findings from Moen et al ^12^, our data shows that the same SNPs that in mothers associate with child’s birth weight exert independent genetic effects on disease risks when transmitted to children. Reflecting this genetic pleiotropy, the disease risks associated with the birth weight scores were consistently associated more closely with the effects from the fetal rather than maternal genome. A clear example of such cases were the CHD and hypertension risks, for which there was consistently very little evidence of any intrauterine effects in play, after taking into account the child PGSs. Our findings thus support the idea that blood pressure and birth weight are connected through the alleles that first reduce the child’s birth weight when present in the mother, and then increase the child’s blood pressure when present in the child, as previously suggested in smaller samples ^15^. Based on our findings, a similar mechanism appears to hold for CHD risk.

Although our data thus allows to conclude that the effects on disease risks mediated by the birth weight PGSs act principally through the child genome, we deteced a couple of instances where the results were less clear-cut. For example, when assessing a child’s risk for receiving statin medication, our data did not explicitly exclude maternal contributions, whereas the results from the father-child pairs pointed to potential post-natal effects. However, the result should be followed up and validated in additional datasets. Finally, our data highlighted associations between birth weight and the risk of T2D, previous studies backing both intrauterine and genetic mechanisms behind this connection ^13,15,26^. Our results from those PGSs that were based on alleles with largely fetal effects on birth weight (BW-68 and BW-201) clearly supported the fetal insuling hypothesis, stating that the same genetic factors that increase birth weight in the fetal genome, also protect from T2D ^6,7^. However, in contrast, the genome-wide PGS for maternal effects in birth weight, in our data rather associated with increased T2D risk in the children. It thus seems that the genetic variants influencing birth weight can have rather complex effects on lifetime T2D risk, depending on their means of action.

Despite the many benefits of using biobank data to explore the connections between maternal traits and child’s disease outcomes, our study and datasets include limitations. In this study, we used genetic variants associated with birth weight as quantitative traits to proxy intrauterine growth restriction in a population-based sample. The birth weight PGSs that were used as instrumental variables in our analyses showed clear effects on birth weight and disease risk at population level.

Hence, we estimate that these are valid instruments to explore the known epidemiological connections between birth weight and disease risks later in life under the MR setting. We nonetheless stress that all the PGSs were based on common genetic variants and explain only a proportion of total variance in birth weight (1SD change in our genetic instrument (MBW-29) corresponded to ∼41g change in birth weight). We thus acknowledge that in this study we may not explicitly model for example severe intrauterine growth restriction resulting from external causes.

Further, instead of reflecting solely intrauterine growth restriction, the PGSs may be partly related to normal variation in child’s size, for example, due to gestational duration, though our results adjusting for the PGS for gestational length suggest that controlling for this has negligible effect on the disease associations (Supplementary Table 5) ^17,27^. In addition, though we used established MR principles, and therefore could test for evidence of a causality of maternal effects on birth weight on disease risk, we have not performed formal MR to provide accurate effect estimates for the effects of birth weight. Also, we assume a simple monotonic relationship between child’s disease risk and the PGS for birth weight, which for some phenotypes can be suboptimal, since both low and high birth weight can increase the risk of same disease ^3^. Finally, our power analyses also indicate that we have likely been limited to detecting maternal effects that are relatively large.

The key medical implication from this work, however, is that modest intrauterine growth restriction during pregnancy is unlikely to have large effects on child’s disease risk in later life. Our findings, aligned with earlier genetic studies, instead support the model that genetic factors in the maternal genome, affecting child’s birth weight, exert independent effects on disease risks later in life when transferred to child genome. Overall, our study demonstrates how MR in genotyped mother-child pairs is a sound and powerful method to evaluate how maternal and fetal exposures relate to child’s health. We envision that large-scale population-based biobanks, such as FinnGen applied here, can boost power for such studies and that these will allow for testing many other hypotheses in the field.

## Methods

### Ethics statement

All patients and control participants in FinnGen provided informed consent for biobank research, based on the Finnish Biobank Act. Research cohorts collected prior the start of FinnGen (in August 2017), were collected based on study-specific consents and later transferred to the Finnish biobanks after approval by Valvira, the National Supervisory Authority for Welfare and Health. Recruitment protocols followed the biobank protocols approved by Valvira. The Coordinating Ethics Committee of the Hospital District of Helsinki and Uusimaa (HUS) approved the FinnGen study protocol Nr HUS/990/2017.

The FinnGen study is approved by Finnish Institute for Health and Welfare (THL), approval number THL/2031/6.02.00/2017, amendments THL/1101/5.05.00/2017, THL/341/6.02.00/2018, THL/2222/6.02.00/2018, THL/283/6.02.00/2019, THL/1721/5.05.00/2019, Digital and population data service agency VRK43431/2017-3, VRK/6909/2018-3, VRK/4415/2019-3 the Social Insurance Institution (KELA) KELA 58/522/2017, KELA 131/522/2018, KELA 70/522/2019, KELA 98/522/2019, and Statistics Finland TK-53-1041-17. The Biobank Access Decisions for FinnGen samples and data utilized in FinnGen Data Freeze 6 include: THL Biobank BB2017_55, BB2017_111, BB2018_19, BB_2018_34, BB_2018_67, BB2018_71, BB2019_7, BB2019_8, BB2019_26, Finnish Red Cross Blood Service Biobank 7.12.2017, Helsinki Biobank HUS/359/2017, Auria Biobank AB17-5154, Biobank Borealis of Northern Finland_2017_1013, Biobank of Eastern Finland 1186/2018, Finnish Clinical Biobank Tampere MH0004, Central Finland Biobank 1-2017, and Terveystalo Biobank STB 2018001.

### FinnGen study

The FinnGen study (https://www.finngen.fi/en) is an on-going research project that utilizes samples from a nationwide network of Finnish biobanks and digital health care data from national health registers ^23^. The goal of the project is to produce genomic data with linkage to health register data for over 500,000 biobank participants nation-wide. The majority of the samples have been gathered from six university hospital biobanks. In the present study, we included samples from 430,897 biobank participants with genotypes available (FinnGen release 10). The samples are linked to national hospital discharge (available from 1968), death (1969–), cancer (1953–) and medication reimbursement (1964–) registries. Additional registries include national birth registry (1987-) containing, e.g., data for birth weight, and the registry on medication purchases (1995-). Currently, after sample pruning and quality control the release 10 of the dataset contains phenotypes for 412,176 participants (181,869 men and 230,307 women, median age of 62.9 yrs), representing roughly 8% of the Finnish population. Due to the sample ascertainment and selection procedures, the cohort has clear advantages over some other population-based sample collections. For example, given that most samples are from hospital biobanks, FinnGen includes an excess of disease cases. However, due to the same reasons FinnGen should not be considered as an epidemiologically representative dataset ^23^.

The parent-offspring relationships (total N=72,465) used in this study had been inferred from the genetic data by the FinnGen analysis team with KING software using the suggested tresholds for calling first degree relatives ^28^. After sample pruning, e.g., excluding duplicate samples and ethnic outliers, and removing suggested parent-child relationships with age difference between samples <15 years, we were left with 67,986 parent-child relationships in the dataset (36,211 mother-child pairs, 31,775 father-child pairs with phenotype data available for analysis). We identified altogether 28,582 unique mothers, with an average of 1.26 children per parent (Supplementary Figure 7). The mean age of the children from the mother-child pairs is 45.0 yrs (SD 14.5).

### Disease endpoints and phenotype data

Birth weight data was available for 39,578 participants in FinnGen R10, born after 1987. For our main analyses, we included the following five predefined endpoints from the FinnGen registry team: coronary heart disease (I9_CHD), hypertension (FG_HYPERTENSION), statin use (RX_STATIN, a proxy for high cholesterol levels), type 2 diabetes (E4_DM2_STRICT) and cancer (C3_CANCER_EXALLC). The disease case number in FinnGen R10 ranged from 46,959 CHD cases to 144,672 statin users (∼11.3% to 35.1% of the dataset). In the mother-child pairs, the N case range for children was from 996 CHD cases to 6150 statin users (corresponding to 2.8% and 16.7% of the dataset), echoing the observation that many, but not all children are old enough to manifest, e.g., cardiac symptoms. More detailed phenotype descriptions and definitions and summary data for these phenotypes for whole FinnGen R10 dataset are available from risteys.finngen.fi.

### Construction of the maternal and fetal polygenic scores for birth weight

We constructed altogether four different polygenic scores (PGSs) to study the relationships between child’s birth weight and later life disease risks in the FinnGen cohort. All PGSs were based on GWAS data from the EGG-consortium ^15^. The EGG-consortium data included summary statistics for GWAS of own (N=321,223) and child birth weight (N=230,069), partitioning the genetic effects into maternal and fetal components.

Two genetic scores were designed to capture specifically maternal effects on birth weight, to proxy intrauterine growth restriction. We first constructed a similar unweighted lead SNP-based PGS as used in Moen et al. (2020) ^12^, by summing up the number of genome-wide significant birth weight increasing alleles per individual. The unweighed score for maternal effects on birth weight (MBW-29) was built based on 32 SNPs identified in GWASs on own and offspring birth weight. Upon partitioning genetic effects into maternal and fetal components using structural equation modelling (SEM), these 32 SNPs were reported to have specifically a maternal effect on birth weight ^15^. Secondly, we calculated a genome-wide polygenic score (PGSs) for maternal effects on birth weight (MBW-GW), based on the summary statistics of a GWAS on offspring birth weight, adjusting for fetal effects using an extension of SEM ^15^.

In addition we constructed two additional scores, of which BW-68 was based on 72 SNPs from the birth weight GWASs, consisting of the 32 SNPs with specifically maternal effects on birth weight, 27 SNPS with directionally concordant maternal and fetal effects on birth weight, and 15 SNPs with directionally opposing maternal and fetal effects ^15^. Finally, we calculated an unweighed score (BW-201) based on the beforementioned 72 SNPs, 64 SNPs with fetal-only effects, and 71 unclassified SNPs ^15^.

The unweighed scores were calculated with plink2 (www.cog-genomics.org/plink/2.0/) ^29^. For the unweighed scores, in FinnGen, we found data for 29, 68 and 201 SNPs respectively (Supplementary Tables 8-10). The relationships of the studied PGSs are illustrated in Supplementary Figure 1. The use of unweighed PGSs has been previously argued to be a more valid measurement for maternal effects on birth weight since the exact effect sizes for the SNPs on the intrauterine growth restriction are unknown ^12^. We chose to also include a genome-wide score based on maternal allelic weights into our study since this had more power to explain variance in birth weight compared to the lead SNP-based scores (Supplementary Table 1). The genome-wide scores were calculated with PRS-CS ^30^ using FinnGen PRS pipeline, filtering data from GWAS of maternal effects on birth weight adjusted for child’s effects to include only HapMap3 SNPs. The downside of using the genome-wide scores, that take the maternal allele weights for both the mother and the child, is that we cannot as accurately control for the pleiotropic effects of the variants in the child’s genome as when using lead SNP-based scores. In theory, relying only on the genome-wide PGSs thus might lead to an excess of false positives (PGS of mother associates with child’s disease through child’s genome rather than through maternal effects during the pregnancy).

We standardized all PGSs into z-scores and reported their effects per SD-unit in our analyses. The EGG-consortium samples based on which the PGSs were built are largely independent of the target samples in the FinnGen, although we note that ∼4.7 % of the EGG consortium GWAS participants are of Finnish ancestry ^15^. We could identify some FinnGen samples that have been included in calculating the GWAS summary statistics for EGG, but these overlapping samples make up only ∼0.4% of the FinnGen mother-child cohort (from NFBC66 and NFBC86, N=146). We consider their potential effects on the results negligible.

### Statistical Analysis

In our primary analyses, we tested for associations between mothers’ PGSs for maternal effects on birth weight (MBW-29) and child’s disease, adjusting for the same PGS computed for the child. The principles of the MR framework are illustrated in Figure 1. The analyses were run using logistic regression (glm function with family =”binomial” in R), and were adjusted for child age, first 10 principal components of genetic structure and genotyping batch. The results from these analyses were obtained as logarithm of odds ratio (logOR) and its standard error (SE) per standard deviation (SD) increase in the PGS. For our main tables we transformed the logOR values and the corresponding SEs or 95% confidence intervals (95%CI) to odds ratio (OR) scale for more intuitive interpretation. All analyses were performed using R Statistical Software (v4.3; R Core Team 2023).

As sensitivity analyses, we performed similar analyses using only one child per mother (N=28,582), in mother-child pairs where 3^th^ degree relatives and closer for both mothers and children had been removed from the analysis, keeping only the oldest mother and her oldest child from an extended family (N=22,454) based on relationships identified in the KING analysis ^28^, and adjusting for polygenic scores for maternal effects on gestational length ^17^. We also utilized the identified father-child pairs (N=31,775) to test for the presence of potential postnatal effects and to further control for potential familial effects. Given the case overlap between most of the disease endpoints and the direct relationships between the PGSs, we did not adjust for multiple testing and we use *p* < 0.05 as the significance threshold.

We acknowledge that some associations between birth weight and disease show a J-shaped curve (both low and very high birth weight increase disease risk compared to more typical birth weight) ^3^. The associations to higher birth weight are anyhow visible only with very high birth weights that are likely outside the variation that our genetic instruments capture, and we therefore chose to include only monotonic effects in our analyses.

### Power calculations

We conducted simulations to estimate the statistical power of our framework to capture maternal effects on child’s endpoints under different combinations of true effects of mother’s and child’s PGSs (Supplementary Code). We ran 1000 simulations for each combination of maternal and child effects on a given endpoint. In each simulation, we:

1. used the mvnorm R package (ref) to generate N=36,211 samples from two-dimensional normal distribution (means 0, variances 1, correlation 0.5) to reflect the birth weight PGSs for the FinnGen mother-child pairs,
2. using these distributions, the case number for the studied endpoint, and chosen maternal and child effects constructed an exponential risk function, an inverse of logistic regression, where the prevalence of the disease matched the observed prevalence in the FinnGen data, and the effect sizes are on a logOR scale.

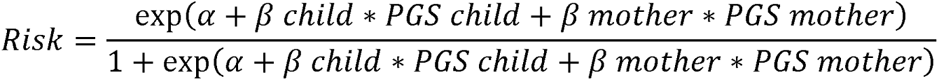
3. randomly sampled a case-control status for each child according to the child’s risk value, and
4. then regressed the sampled case-control vector jointly on the mother’s and child’s PGSs using logistic regression.

The estimate for power was obtained as the proportion of simulations where the p-value for the coefficient of interest (mother’s or child’s PGS) was below the p-value threshold of 0.05. Power calculations were conducted in R (version 4.2.3 (2023-03-15)).

## Data Availability

Full genetic and clinical data from FinnGen is available for researchers by application (https://www.finngen.fi/en/access_results). Details of FinnGen core endpoints can be found at risteys.finngen.fi. The source data for results figures is available in Supplementary Tables: for Figure 2 - Supplementary Table 4; Supplementary Figure 3 – Supplementary Table 4; Figure 4 – Supplementary Tables 4, 5 and 6; Figure 5 – Supplementary Data 6.

## Code availability

The code for power analyses is available as Supplementary Code. The full genotyping and imputation protocol for FinnGen is described at https://doi.org/10.17504/protocols.io.nmndc5e.

## Acknowledgements

We thank all FinnGen participants, principal investigators, laboratory personnel and data management teams. The FinnGen project is funded by two grants from Business Finland (HUS 4685/31/2016 and UH 4386/31/2016) and the following industry partners: AbbVie Inc., AstraZeneca UK Ltd, Biogen MA Inc., Bristol Myers Squibb (and Celgene Corporation & Celgene International II Sàrl), Genentech Inc., Merck Sharp & Dohme LCC, Pfizer Inc., GlaxoSmithKline Intellectual Property Development Ltd., Sanofi US Services Inc., Maze Therapeutics Inc., Janssen Biotech Inc, Novartis AG, and Boehringer Ingelheim International GmbH. Following biobanks are acknowledged for delivering biobank samples to FinnGen: Auria Biobank (www.auria.fi/biopankki), THL Biobank (www.thl.fi/biobank), Helsinki Biobank (www.helsinginbiopankki.fi), Biobank Borealis of Northern Finland (https://www.ppshp.fi/Tutkimus-ja-opetus/Biopankki/Pages/Biobank-Borealis-briefly-in-English.aspx), Finnish Clinical Biobank Tampere (www.tays.fi/en-US/Research_and_development/Finnish_Clinical_Biobank_Tampere), Biobank of Eastern Finland (www.ita-suomenbiopankki.fi/en), Central Finland Biobank (www.ksshp.fi/fi-FI/Potilaalle/Biopankki), Finnish Red Cross Blood Service Biobank (www.veripalvelu.fi/verenluovutus/biopankkitoiminta), Terveystalo Biobank (www.terveystalo.com/fi/Yritystietoa/Terveystalo-Biopankki/Biopankki/) and Arctic Biobank (https://www.oulu.fi/en/university/faculties-and-units/faculty-medicine/northern-finland-birth-cohorts-and-arctic-biobank). All Finnish Biobanks are members of BBMRI.fi infrastructure (www.bbmri.fi). Finnish Biobank Cooperative-FINBB (https://finbb.fi/) is the coordinator of BBMRI-ERIC operations in Finland. The Finnish biobank data can be accessed through the Fingenious^®^ services (https://site.fingenious.fi/en/) managed by FINBB.

T.T. was funded by the Research Council of Finland (grant numbers 315589 and 345867), Sigrid Jusélius Foundation (https://sigridjuselius.fi/en/), and the HiLIFE Fellows Program. M.P. was supported by the Research Council of Finland (https://www.aka.fi/en/) grant 338507, Research Council of Finland Center of Excellence in Complex Disease Genetics grant 352795, and the Sigrid Juselius Foundation.

## Author Contributions

JL and TT conceived the study. JL and TT performed data analyses. TT formulated the power analyses. MP provided statistical support. JL and TT drafted to original manuscript. JL, MP and TT critically revised the draft manuscript. All authors approved the final version of the manuscript.

## Competing interests

The authors declare no competing interests

## Notes

### Competing Interest Statement

The authors have declared no competing interest.

### Author Declarations

All patients and control participants in FinnGen provided informed consent for biobank research, based on the Finnish Biobank Act. Research cohorts collected prior the start of FinnGen (in August 2017), were collected based on study-specific consents and later transferred to the Finnish biobanks after approval by Valvira, the National Supervisory Authority for Welfare and Health. Recruitment protocols followed the biobank protocols approved by Valvira. The Coordinating Ethics Committee of the Hospital District of Helsinki and Uusimaa (HUS) approved the FinnGen study protocol Nr HUS/990/2017. The FinnGen study is approved by Finnish Institute for Health and Welfare (THL), approval number THL/2031/6.02.00/2017, amendments THL/1101/5.05.00/2017, THL/341/6.02.00/2018, THL/2222/6.02.00/2018, THL/283/6.02.00/2019, THL/1721/5.05.00/2019, Digital and population data service agency VRK43431/2017-3, VRK/6909/2018-3, VRK/4415/2019-3 the Social Insurance Institution (KELA) KELA 58/522/2017, KELA 131/522/2018, KELA 70/522/2019, KELA 98/522/2019, and Statistics Finland TK-53-1041-17. The Biobank Access Decisions for FinnGen samples and data utilized in FinnGen Data Freeze 10 include: THL Biobank BB2017_55, BB2017_111, BB2018_19, BB_2018_34, BB_2018_67, BB2018_71, BB2019_7, BB2019_8, BB2019_26, BB2020_1, BB2021_65, Finnish Red Cross Blood Service Biobank 7.12.2017, Helsinki Biobank HUS/359/2017, HUS/248/2020, HUS/150/2022 12, 13, 14, 15, 16, 17, 18, and 23, Auria Biobank AB17-5154 and amendment #1 (August 17 2020) and amendments BB_2021-0140, BB_2021-0156 (August 26 2021, Feb 2 2022), BB_2021-0169, BB_2021-0179, BB_2021-0161, AB20-5926 and amendment #1 (April 23 2020)and it's modification (Sep 22 2021), Biobank Borealis of Northern Finland_2017_1013, 2021_5010, 2021_5018, 2021_5015, 2021_5023, 2021_5017, 2022_6001, Biobank of Eastern Finland 1186/2018 and amendment 22/2020, 53/2021, 13/2022, 14/2022, 15/2022, Finnish Clinical Biobank Tampere MH0004 and amendments (21.02.2020 & 06.10.2020), 8/2021, 9/2022, 10/2022, 12/2022, 20/2022, 21/2022, 22/2022, 23/2022, Central Finland Biobank 1-2017, and Terveystalo Biobank STB 2018001 and amendment 25th Aug 2020, Finnish Hematological Registry and Clinical Biobank decision 18th June 2021, Arctic biobank P0844: ARC_2021_1001.

