## Supplementary Figures 1-7 and Supplementary Code for "Disentangling the link between maternal influences on birth weight and disease risk in 36,211 genotyped mother-child pairs"

### Supplementary information

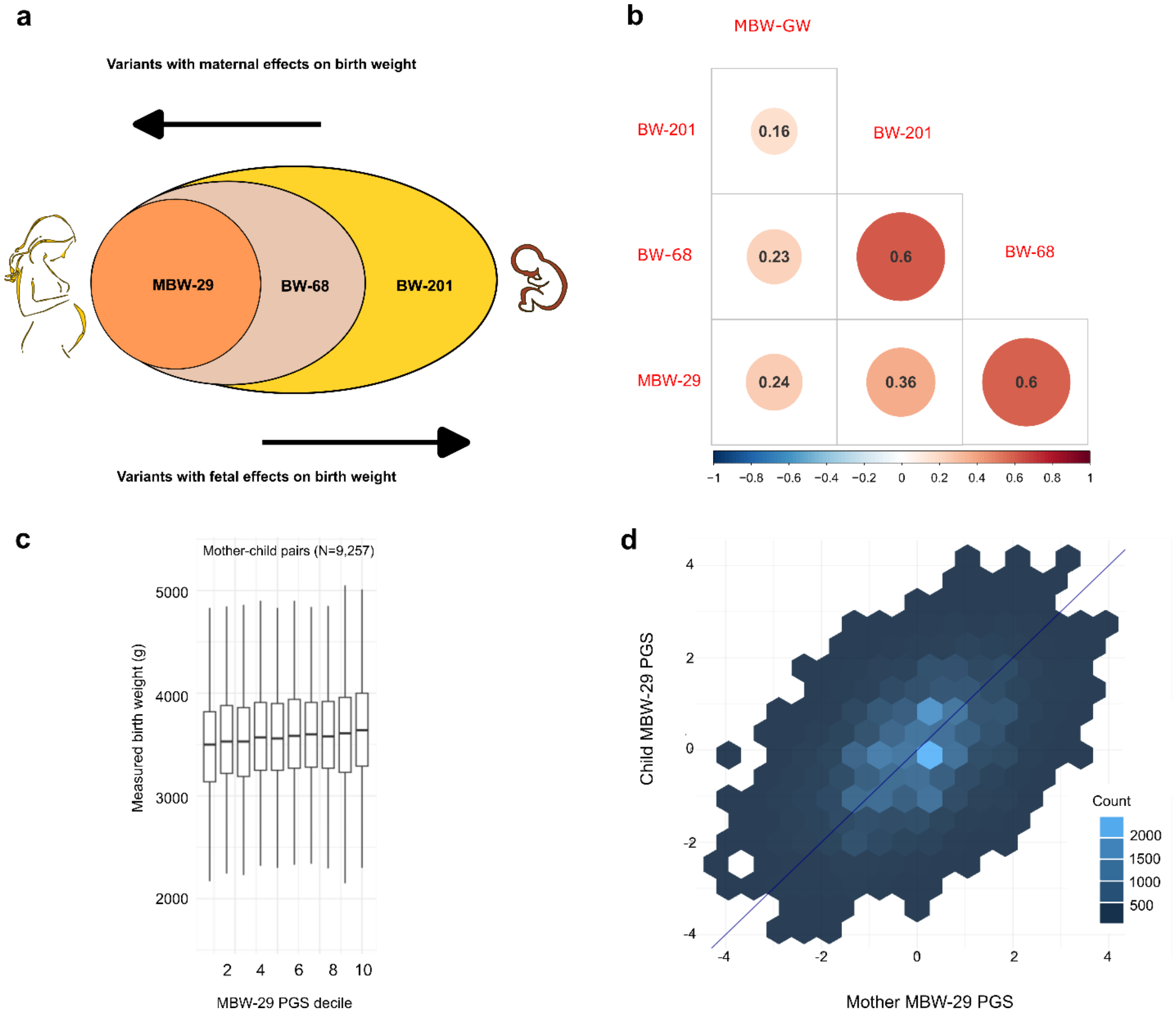


Supplementary Figure 1. Schematic illustration of the relationships between the studied PGSs, their correlations, and their effects. Panel a) illustrates the relationships between the unweighed lead-SNP based PGSs (MBW-29, BW-68 and BW-201) which partly overlap with each other in the illustrated manner. MBW-29 is solely based on variants with maternal effects on birth weight only, BW-68 is based on variants that have maternal effects on birth weight, some of which have also fetal effects, and BW-201 is mostly based on SNPs that act through the child's genome to affect birth weight, though it contains also the maternal SNPs. b) The correlations between the PGSs, including also the genome-wide score for maternal effects on birth weight (MBW-GW). c) The effect of MBW-29 PGS on child's birth weight in mother-child pairs. d) Illustration of the correlation (pearson r = 0.504) of mother's and child's MBW-29 PGS in the FinnGen mother-child pairs. Each hexagon is based on data from at least 15 mother-child pairs. Lighter blue shades indicate more mother-child pairs under each hexagonal bin.

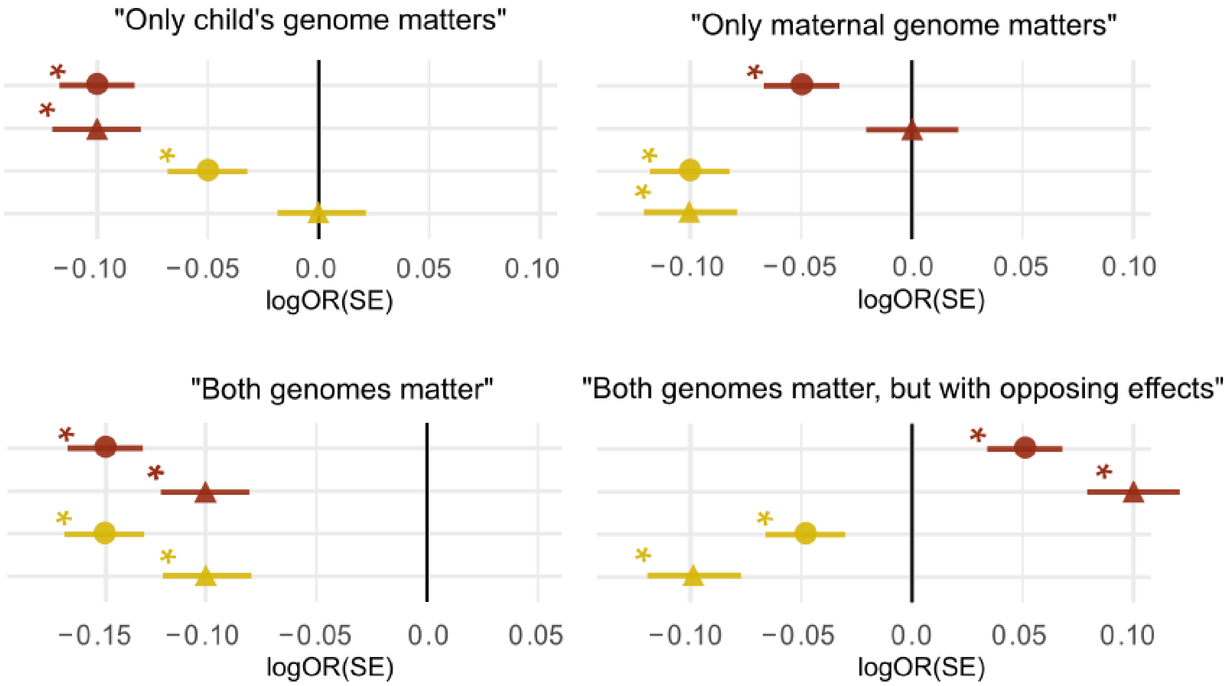


Supplementary Figure 2. Simulations showing how we would expect the PGS associations to look like under four different scenarios (1) only child genome matters, 2) only maternal genome matters, 3) both genomes matter and 4) both genomes matter, exerting opposite effects. The simulations are based on true effect size of logOR -0.1 and N cases =4,700. Red = effect estimates for the child's PGS. Yellow = effect estimates for the mother's PGS. Round dots = effect from a model where only mother's or child's PGS has been included into the analysis. Triangles = effect from a model where both mother's and child's PGS have been included into the same analysis.

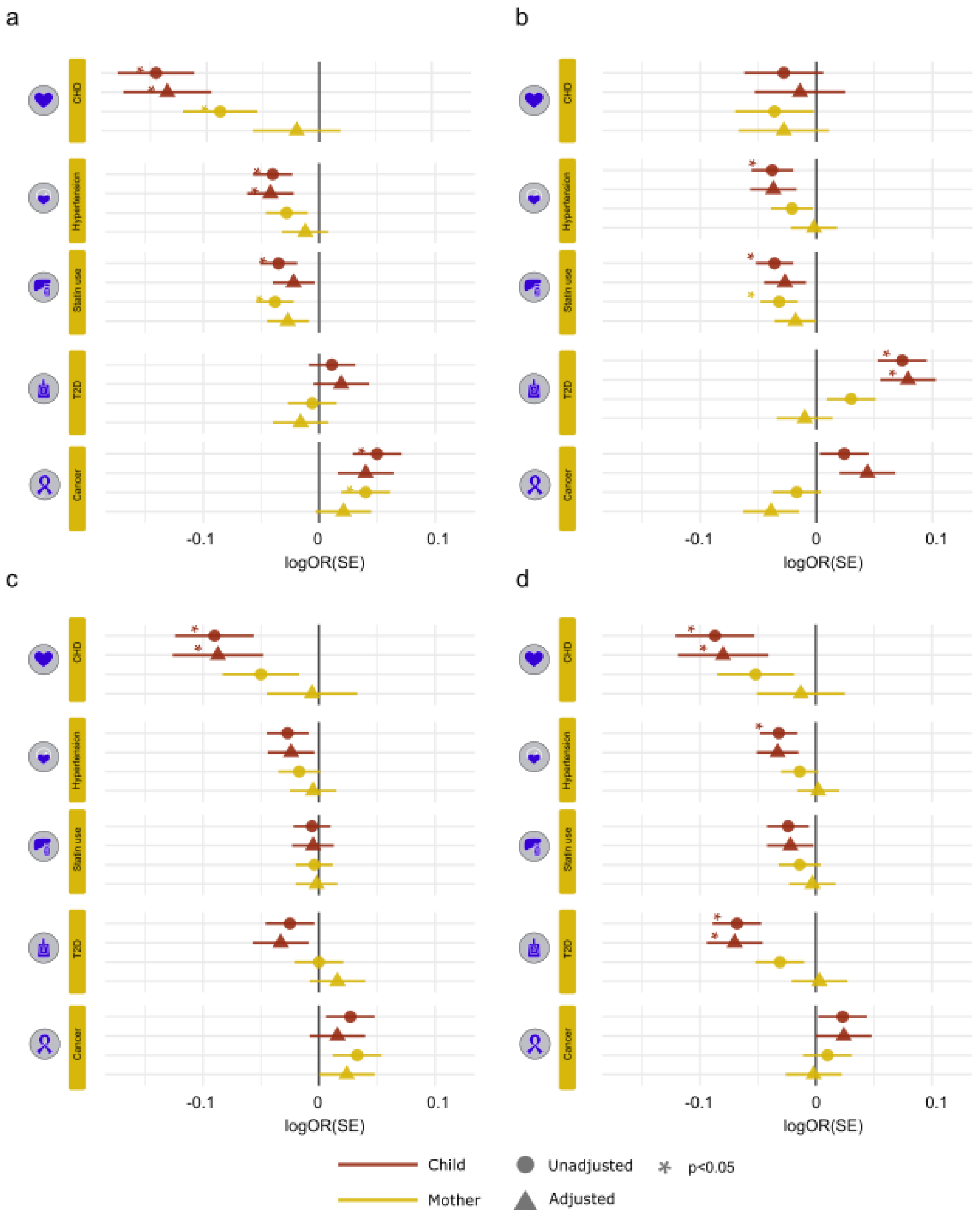


**Supplementary Figure 3. Associations of PGSs MBW-29, MBW-GW, BW-68 and BW-201 with disease risk in the children from mother-child pairs (N=36,211)**. The figures shows how the child's own PGS (red) and mother's PGS (yellow) associate with child's disease risk in the mother-child pairs. a) Effect of 1SD increase in MBW-29 on disease risk.b) Effect of 1SD increase in MBW-GW on disease risk. c) Effect of 1SD increase in BW-68 on disease risk. d) Effect of 1SD increase in BW-201 on disease risk. Round dots illustrate cases when only one PGS is used in the analyses, and triangels indicate that the analyses include PGSs from both mothers and their children. The lines mark standard error (SE). * denotes statistically significant association between a PGS and disease risk in children.

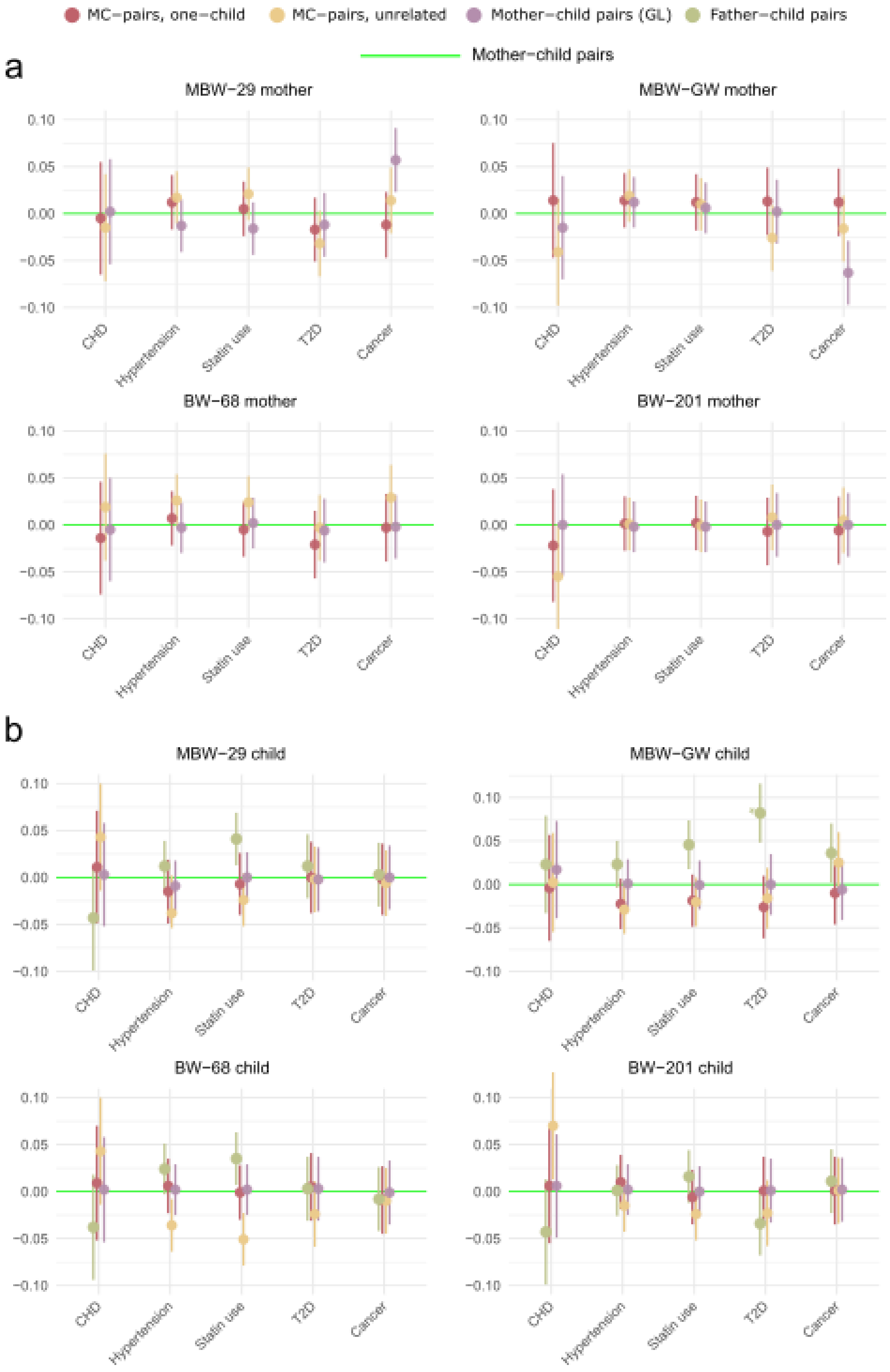


**Supplementary Figure 4. Change in effect sizes in logOR units between different models.** Dots indicate point estimates with lines showing error bars (SE). Green line refers to effect in the main analyses in mother-child pairs (mother and child PGS effect on disease taking into account both PGSs). Red=effect difference in mother-child pairs where only one child has been included, yellow= effect difference in unrelated pairs, lilac= effect difference in mother-child pairs adjusting for PGS for maternal effects on gestational length (GL), green = the effect difference for children in the father-child pairs. The asterisk marks statistically significant difference in effect sizes between the two models.

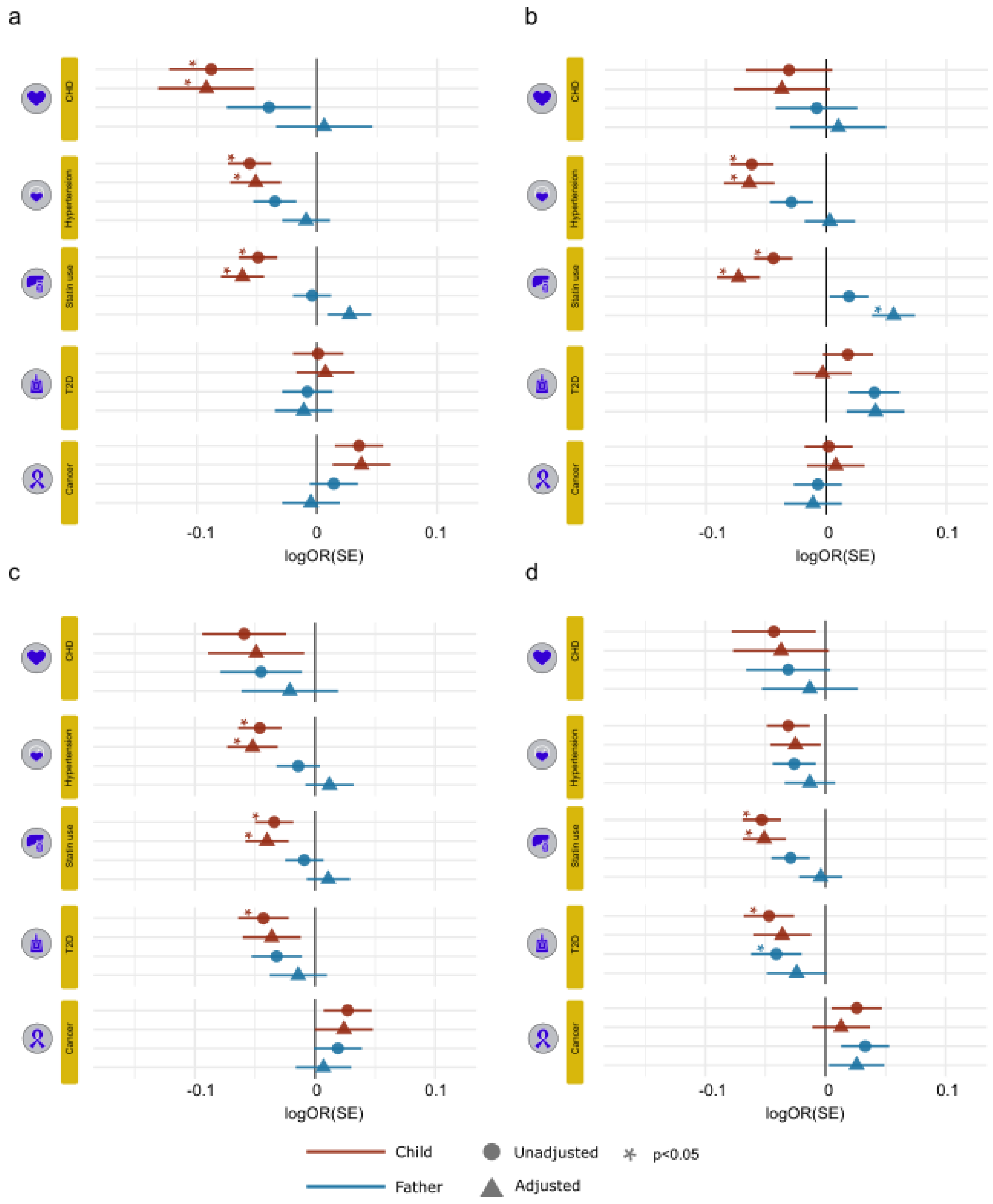


**Supplementary Figure 5. Associations of PGSs MBW-29, MBW-GW, BW-68 and BW-201 with disease risk in the children from father-child pairs (N=31,775)**. The figures shows how the child's own PGS (red) and father's PGS (blue) associate with child's disease risk in the father-child pairs. a) Effect of 1SD increase in MBW-29 on disease risk. b) Effect of 1SD increase in MBW-GW on disease risk. c) Effect of 1SD increase in BW-68 on disease risk. d) Effect of 1SD increase in BW-201 on disease risk. Round dots illustrate cases when only one PGS is used in the analyses, and triangels indicate that the analyses include PGSs from both fathers and their children. The lines mark standard error (SE). * denotes statistically significant association between a PGS and disease risk in children.

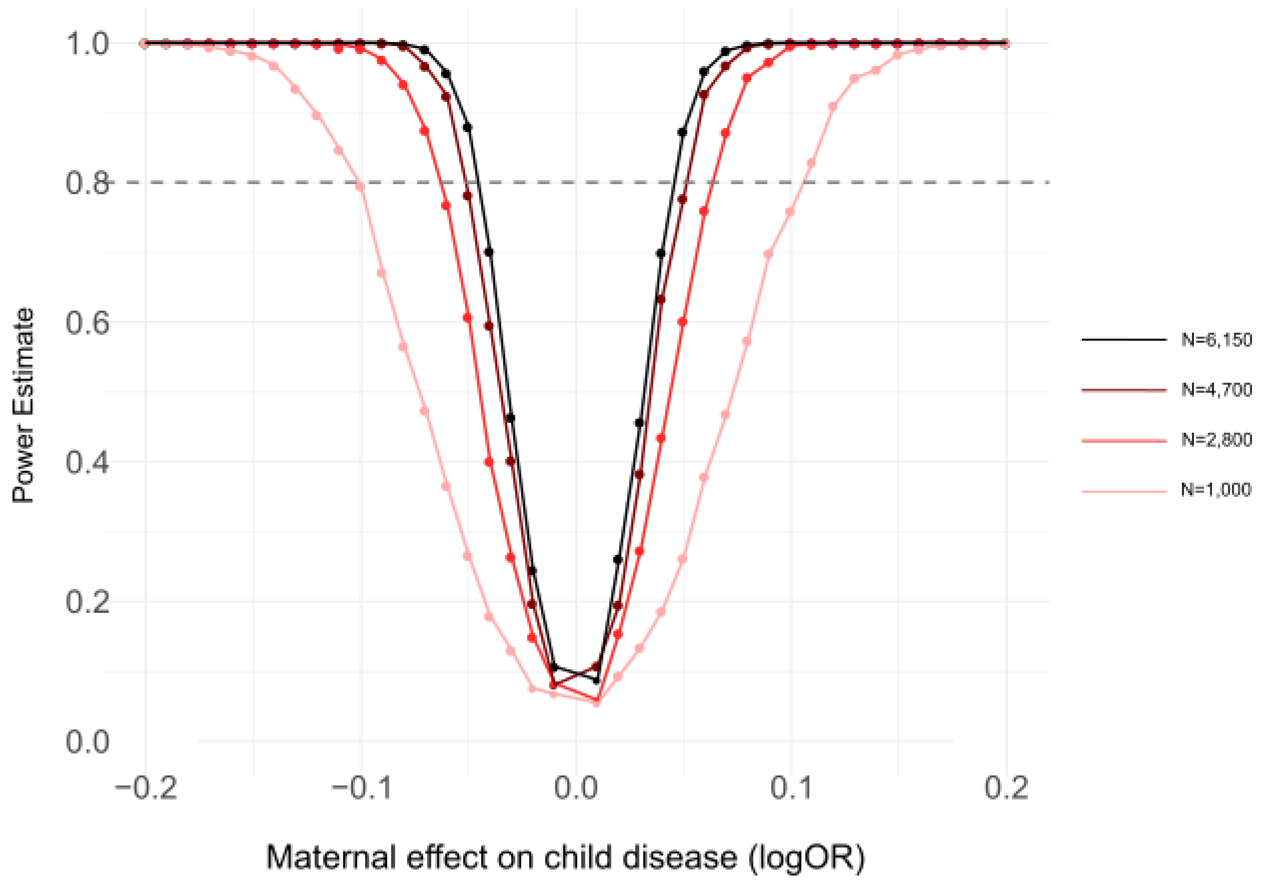


**Supplementary Figure 6. Illustration of power to detect maternal effects on child disease depending on number of cases in the mother-child pairs (N=36,211),** The point estimates and fitted curves illustrate power when taking into account both child and mother effects simultaneously (Supplementary Table 3). The estimates are based on assumed true maternal effects of logOR -0.2 to 0.2 in 0.01 unit intervals in mothers, and no effects in children. Each data point is based on 1000 simulations. Colours indicate number of disease cases in each simulation in 36,211 mother-child pairs. Black = N cases 6,150, dark red N = 4,700, red N =2,800, pink N = 1000, corresponding to observed disease cases in FinnGen mother-child pairs for statin use, hypertension, T2D and cancer, and CHD, respectively.

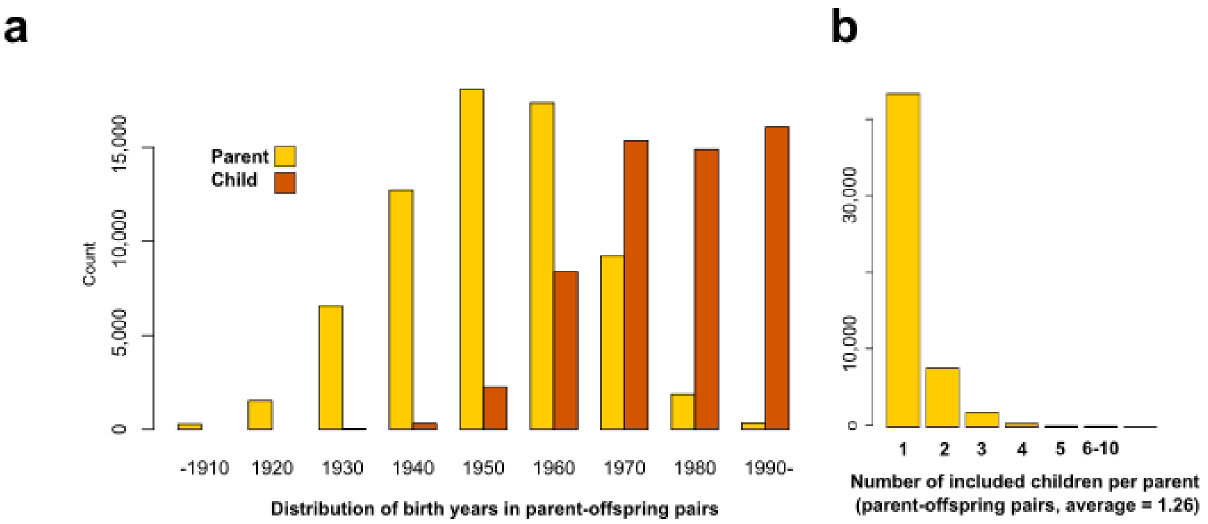


Supplementary Figure 7. a Distribution of the birth years of the parent-offspring pairs in FinnGen. b Number of children per parent in the data set.

**Supplementary code.** Simulate power for binary endpoints in the mother-child pairs.

```
sim_pgs_power <- function(Nmc,Ncases,betao,betam){

library(mvtnorm)

### Simulate mother and child polygenic scores (PGS)

# Covariance matrix for maternal and offspring PGS (r=0.5) scaled to unit variance

sigma <- matrix(c(1,0.5,0.5,1),nrow=2,byrow=T)

# Simulate PGS distributions

mc_pgs <- rmvnorm(Nmc,mean=c(0,0),sigma,method="eigen")

### Construct phenotype

# Set alpha and risk function

k = Ncases/Nmc

alpha = log(k/(1-k))

risk <- exp(alpha+betao*mc_pgs[,1]+betam*mc_pgs[,2])/(1+exp(alpha+betao*mc_pgs[,1]+betam*mc_pgs[,2]))

### Test power

# Case-control vector

cco <- rbinom(Nmc,size=1,risk)

# Generalised linear regression: case status ~ offspring PGS + maternal PGS

out <- summary(glm(cco~mc_pgs[,1]+mc_pgs[,2],family='binomial'))$coefficients

# Output results: intercept, offspring beta, maternal beta, offspring p-value, maternal p-value

sim_res <- data.frame(alpha=out[1,1],betao=out[2,1],betam=out[3,1],pbetao=out[2,4],pbetam=out[3,4]

return(sim_res)

}
```
